# Prior reproductive and non-reproductive depression, and depressive symptoms in menopausal transition

**DOI:** 10.64898/2026.06.01.26354583

**Authors:** Marijn Schipper, Margot W.L. Morssinkhof, Dorenda K.E. van Dijken, Yadira Roggeveen, Birit F.P. Broekman

## Abstract

**Importance:** The menopausal transition is associated with an increased risk of depression. Prior depression is a well-established risk factor, but studies do not distinguish between prior reproductive and non-reproductive depression.

**Objective:** To compare the associations of reproductive (i.e., premenstrual mood disorder and perinatal depression) and non-reproductive (i.e., not related to hormonal transitions) histories of depression with depressive symptoms during the menopausal transition.

**Design:** Cross-sectional analysis of questionnaire data from the Multidisciplinary Menopausal Outpatient Care Project (MOPP) collected between February 2023 and October 2025.

**Setting:** Menopause outpatient clinics Amsterdam, the Netherlands, including a specialized multidisciplinary menopause clinic.

**Participants:** In total 364 individuals were approached; 244 enrolled at baseline. After exclusions for age <40 (n=3), premature ovarian insufficiency (n=2), premenopausal status (n=1), age >58 with final menstruation >10 years earlier (n=12), bipolar disorder (n=5), and missing survey data (n=41), 180 participants were included.

**Exposures:** Premenstrual mood disorder measured with Premenstrual Symptom Screening Tool, perinatal depression with Edinburgh Postnatal Depression Scale Lifetime version, and reported prior non-reproductive depression in medical records.

**Main outcome and measures:** Depressive symptom severity measured with Inventory of Depressive Symptomatology-Self Rated. We used univariable and multivariable linear regressions; multivariable models accounted for overlap between exposures.

**Results:** Among 180 participants (median age 51; 61% perimenopausal and 39% postmenopausal), premenstrual mood disorder showed the strongest association with depressive symptom severity (B = 9.0, 95% CI 5.1–12.9, p < 0.001), followed by perinatal depression (B = 7.8, 95% CI 3.4–12.1, p < 0.001) and prior non-reproductive depression (B = 4.7, 95% CI 0.7–8.7, p = 0.021). In multivariable analysis, only premenstrual mood disorder (B = 7.2, 95% CI 2.4–12.1, p = 0.0037) and perinatal depression (B = 5.7, 95% CI 1.2–10.1, p = 0.013) remained associated with depressive symptom severity.

**Conclusions and Relevance:** Prior reproductive depression, but not prior non-reproductive depression, was associated with greater depressive symptom severity during the menopausal transition. A history of premenstrual mood disorder and/or perinatal depression may therefore help identify individuals at increased vulnerability to depressive symptoms during this period. Future studies should replicate these findings in population-based samples.

**Key points:** *Question:* Are histories of prior reproductive depression (i.e., premenstrual mood disorder or perinatal depression) and non-reproductive depression (i.e., not related to hormonal transitions) differentially associated with depressive symptoms during the menopausal transition?

*Findings:* In this cross-sectional study of 180 perimenopausal and postmenopausal patients attending menopause outpatient clinics, prior reproductive depression, but not prior non-reproductive depression, was associated with greater depressive symptom severity during the menopausal transition.

*Meaning:* A history of reproductive depression may help identify individuals at increased risk for depressive symptoms during the menopausal transition.

**Graphical abstract:** 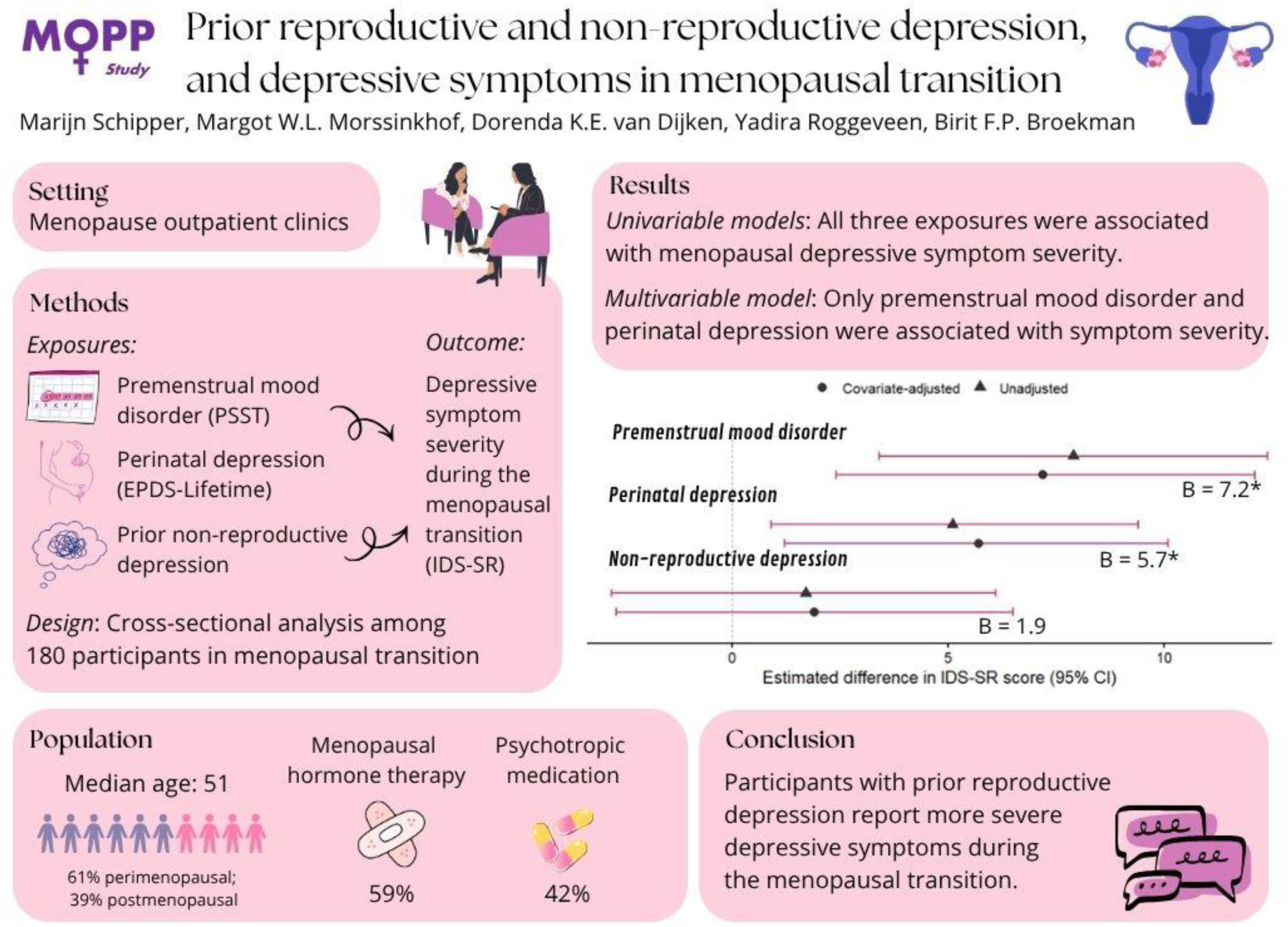

## Introduction

Reproductive transitions are periods of increased vulnerability to depression, including the menopausal transition.^1,2^ While psychosocial factors (i.e., socio-economic adversity, stressful life events, caregiving burden, early trauma, certain personality traits) may influence this increased risk depression during the menopausal transition^1,3–7^, fluctuating reproductive hormone levels could be a key part of the etiology of depression.

The period around the final menstrual period is defined by strong changes in reproductive hormones, across several menopausal stages. The early perimenopause involves great fluctuations in estradiol and progesterone levels, whereas late perimenopause and early postmenopause involve sustained declines in estradiol and progesterone with cessation of menstrual cycles.^8^ Estradiol and progesterone receptors are widely distributed across brain regions that regulate affect, stress responses, and reward processing, and fluctuations in reproductive hormones could therefore affect neurobiological systems involved in mood regulation.^9,10^

Some individuals seem particularly sensitive to the effects of female reproductive hormonal changes on mood.^11–14^ Premenstrual dysphoric disorder (PMDD), characterized by severe mood symptoms during the luteal phase and affecting an estimated 5 to 8% of menstruating individuals, seems to be caused by heightened sensitivity to normal hormonal fluctuations rather than abnormal hormone levels.^15,16^ This has led to the concept of hormonal mood sensitivity, a transdiagnostic vulnerability that may link depressive episodes across reproductive transitions, including PMDD, postpartum depression, and perimenopausal depression.^11–14^ Supporting this idea, a history of PMDD has been associated with postpartum depression^17–20^ and with more severe perimenopausal depressive symptoms^21^. In contrast, evidence linking postpartum depression to perimenopausal depression remains limited. One large registry study found no increased risk among women with postpartum depression compared to women with non-postpartum depression, but they relied on age rather than menopausal stage, likely causing misclassification.^23^

A history of depression is the most consistently identified risk factor for depression during perimenopause and early postmenopause.^4,5,24^ However, studies have rarely distinguished reproductive depression (including PMDD or perinatal depression), from non-reproductive depression (defined as depressive episodes not related to hormonal transitions). It therefore remains unclear whether prior reproductive depression is differently associated with depressive symptoms during the menopausal transition than prior non-reproductive depression. Clarifying these associations may improve the identification of individuals at increased risk and inform more personalized prevention and treatment.

Thus, this study aimed to examine whether prior reproductive and non-reproductive depression are differentially associated with depressive symptom severity during the menopausal transition in a cohort of individuals attending menopause outpatient clinics.

## Methods

### Design

This cross-sectional study used baseline data collected between February 2023 and October 2025 from the Multidisciplinary Menopausal Outpatient Care Project (MOPP), a prospective cohort on mental wellbeing during the menopausal transition at OLVG, a large teaching hospital in Amsterdam, the Netherlands. To ensure sufficient representation of participants with clinically relevant depressive symptoms, participants were recruited from menopause outpatient clinics, including standard gynecology clinics and a multidisciplinary gynecology and psychiatry clinic specialized in the interaction between menopause and psychiatric symptoms. The Medical Ethics Committee United confirmed that the study was not subject to the Dutch Medical Research Involving Human Subjects Act (AW23.029/W22.199). The study was approved by the Advisory Committee for Scientific Research (ACWO) of OLVG hospital (22.123). All participants provided informed consent.

### Participants

Eligible participants were individuals assigned female at birth referred to a menopause outpatient clinic, who could read and write Dutch or English. Study information was sent to 364 individuals, of whom 244 enrolled at baseline. Exclusion criteria were age <40 years (n=3), premature ovarian insufficiency (n=2), classification as premenopausal (n=1), age >58 years with final menstruation >10 years earlier (n=12), and bipolar disorder (n=5). Bipolar disorder was excluded due to distinct pathophysiology compared to unipolar depression and potential hypomanic symptoms not assessed. After excluding 41 participants with missing IDS-SR and/or PSST data, 180 participants were included in the analyses (Figure 1).

**Figure 1.**
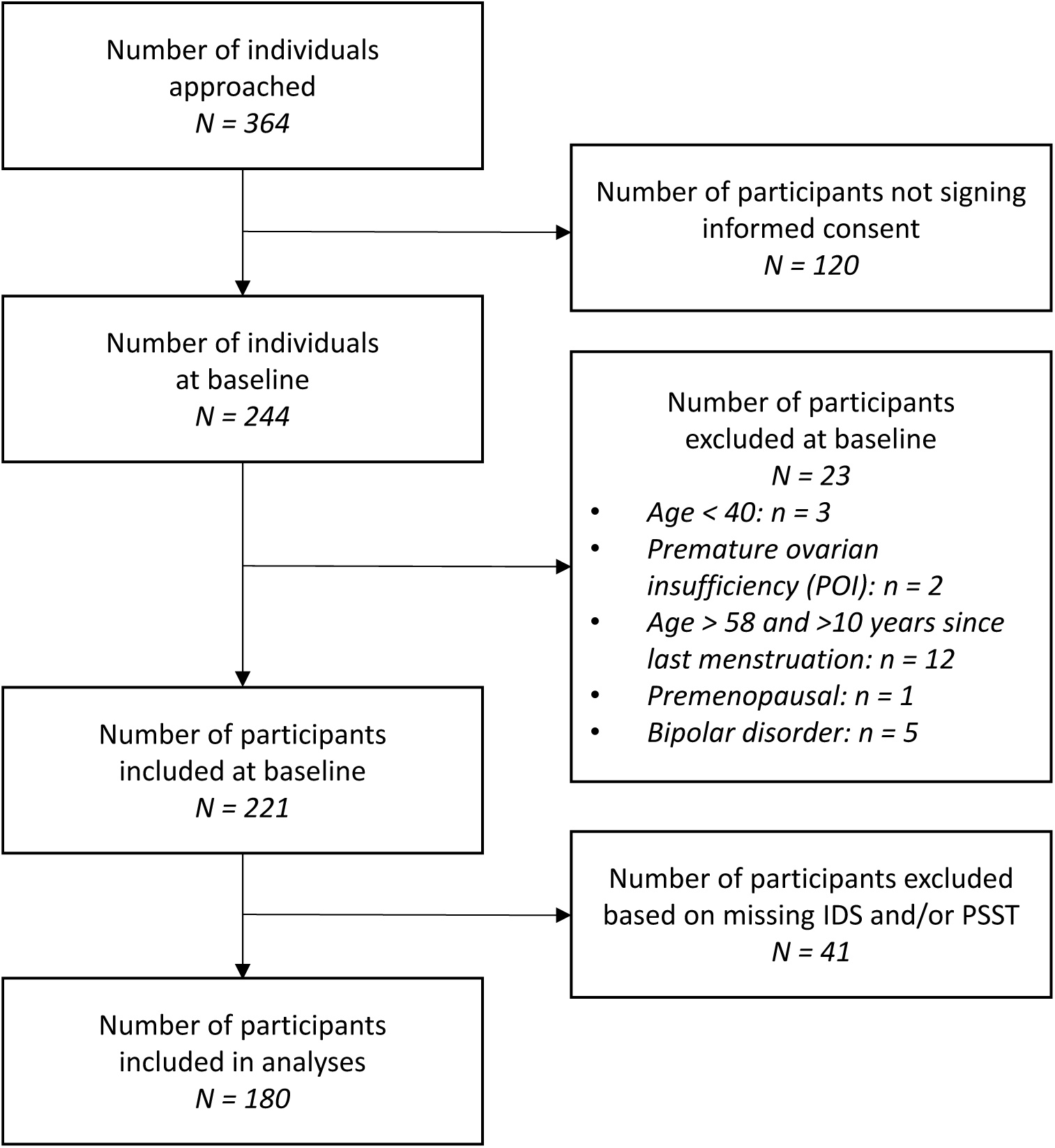
Flow diagram of participant inclusion.

### Procedure

After the first consultation, participants completed online questionnaires (median 15 days later, IQR 6.0-28.0). Clinical data from medical records included age, menopausal status, referring physician, outpatient clinic type, lifestyle factors, and current medication use. Menopausal status at intake was classified according to STRAW criteria, with postmenopause defined as absence of menstruation for more than one year.^8^ Menopausal hormone therapy (MHT) was defined as systemic estrogen therapy, with or without a progestogen, and oral contraceptive as estrogen-containing hormonal contraceptives; used in the week preceding completion of the surveys.

### Outcome: Severity of depressive symptoms

We assessed depressive symptom severity with the 30-item self-rated Inventory of Depressive Symptomatology (IDS-SR), which measures symptoms over the past seven days.^25^ The items of the IDS-SR were summed to a sum score (range: 0-90), with higher scores indicating greater symptom severity: no or minimal (0-13), mild (14-25), moderate (26-38), severe (39-48), and very severe (≥49). The IDS-SR has strong psychometric properties (Cronbach’s α ranging from 0.92 to 0.94 across different populations).^26,27^ Internal consistency was good in the present sample (Cronbach’s α = 0.89).

### Exposures: History of reproductive and non-reproductive depression

We assessed premenstrual mood disorder with the Premenstrual Symptoms Screening Tool (PSST).^28^ Because many participants had irregular or absent cycles, PSST scores were interpreted as reflecting a history of menstrual cycle-related mood symptoms rather than current symptoms. The PSST generally has good internal consistency (Cronbach’s α 0.89-0.93), but lower specificity than prospective daily rating methods.^29–33^ Therefore, we used the PSST as an indicator of premenstrual mood disorder history rather than as a diagnostic assessment of premenstrual mood disorder (PMS) or premenstrual dysphoric disorder (PMDD). Internal consistency of the PSST was excellent in the present sample (Cronbach’s α = 0.95). Following established criteria, we classified participants as having no/mild PMS, moderate/severe PMS, or PMDD.^28^ For analyses, we combined moderate/severe PMS and PMDD into one category representing premenstrual mood disorder.

We assessed perinatal depression using the lifetime version of the Edinburgh Postnatal Depression Scale (EPDS-Lifetime), which captures the lifetime occurrence, timing, and severity of perinatal depressive symptoms.^34^ The EPDS is a widely used screening instrument with strong psychometric properties.^35,36^ Internal consistency was good in the present sample (Cronbach’s α = 0.85). The items were summed (range: 0-30), with scores ≥12 considered indicative of past perinatal depression.^34,35^ For nulliparous participants, EPDS-Lifetime scores were coded as missing, as presence of perinatal depression could not be assessed. Based on symptom timing, episodes were classified as prenatal (during pregnancy) or postpartum (within the first six months after birth).

We assessed non-reproductive depression using psychiatric information in the medical records. At intake, participants reported any prior depressive episodes, including diagnoses and treatments. Clinicians also asked whether symptoms were related to the menstrual cycle or pregnancy, allowing classifications of episodes as non-reproductive depression. Participants without a recorded diagnosis but with strong and consistent clinical indicators, such as long-term antidepressant use, were classified as having probable depression (n=10). All classifications were reviewed by a psychiatrist.

### Covariates

We selected covariates following principles of covariate control described by Van der Weele ^37^. Based on literature, we adjusted the analyses for age^38^, alcohol use and smoking status^39^, MHT use^40–42^, and oral contraceptive use^43^.

### Statistical analyses

Continuous variables were summarized as means with standard deviations (SD) or medians with interquartile ranges (IQR), depending on distribution. Categorical variables were summarized as counts and percentages.

We used linear regression to examine the associations of premenstrual mood disorder, perinatal depression, and prior non-reproductive depression with current depressive symptom severity (IDS-SR score). The distribution of IDS-SR scores was approximately normal; therefore, no transformation was applied. We first fitted separate models for each exposure and then a multivariable model including all three exposures to account for overlap between exposures. Multivariable analyses were restricted to parous participants (live births ≥ 1), as susceptibility to perinatal depression could not be assessed in nulliparous participants.^17–20^ All models were adjusted for age, alcohol use, smoking status, MHT use, and oral contraceptive use.

Results were reported as unstandardized beta coefficients with 95% confidence intervals. Model assumptions were checked and met. Model fit was assessed using log-likelihood and R². Statistical significance was set at p < 0.05. Analyses were conducted in RStudio.^44^

We conducted sensitivity analyses to assess robustness (Supplementary materials). First, we examined the odds of current depression using logistic regression (IDS-SR ≥26 indicating moderate to severe depression). Second, we repeated the analyses in perimenopausal participants only (excluding postmenopausal participants), because perimenopause involves the greatest hormonal fluctuations and depression risk does not appear to remain consistently elevated in postmenopause.^2^ Third, we repeated the analyses using postpartum depression instead of perinatal depression as the exposure, because the postpartum period involves a marked decline in reproductive hormones which may be more comparable to the hormonal changes of the menopausal transition than pregnancy.

## Results

### Demographic characteristics

Median age in our sample was 51 years (IQR 47–54). Of participants, 110 were perimenopausal (61%) and 70 postmenopausal (39%). Most were referred by their general practitioner (n=142, 79%) and attended the standard menopause outpatient clinic (n=157, 87%). Psychotropic medication use was common (n=76, 42%), most often antidepressants (n=49, 27%) or benzodiazepines (n=36, 20%). MHT was used by 106 participants (59%) and oral contraceptives by 16 (9%). Full characteristics are shown in Table 1.

**Table 1.**
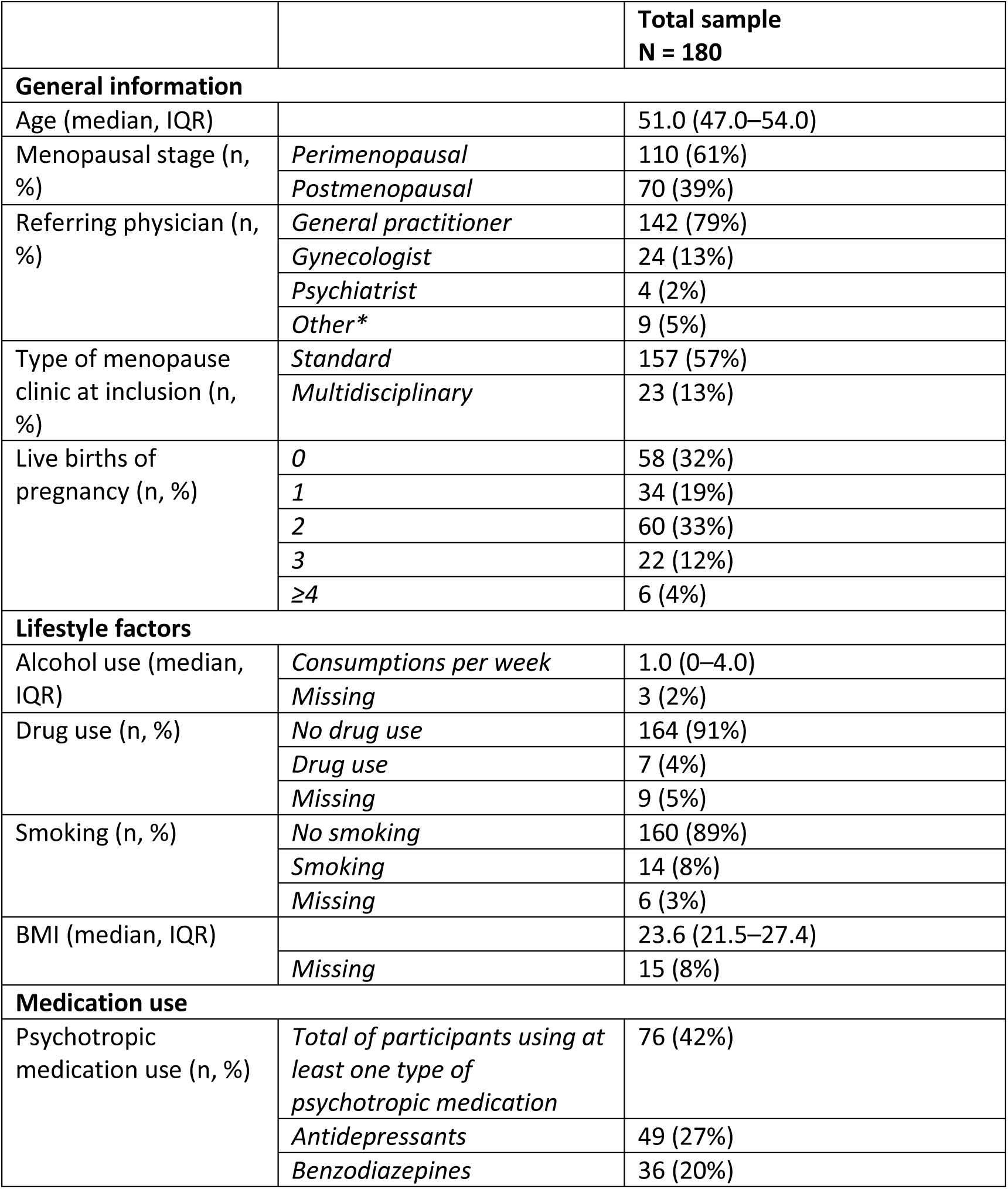
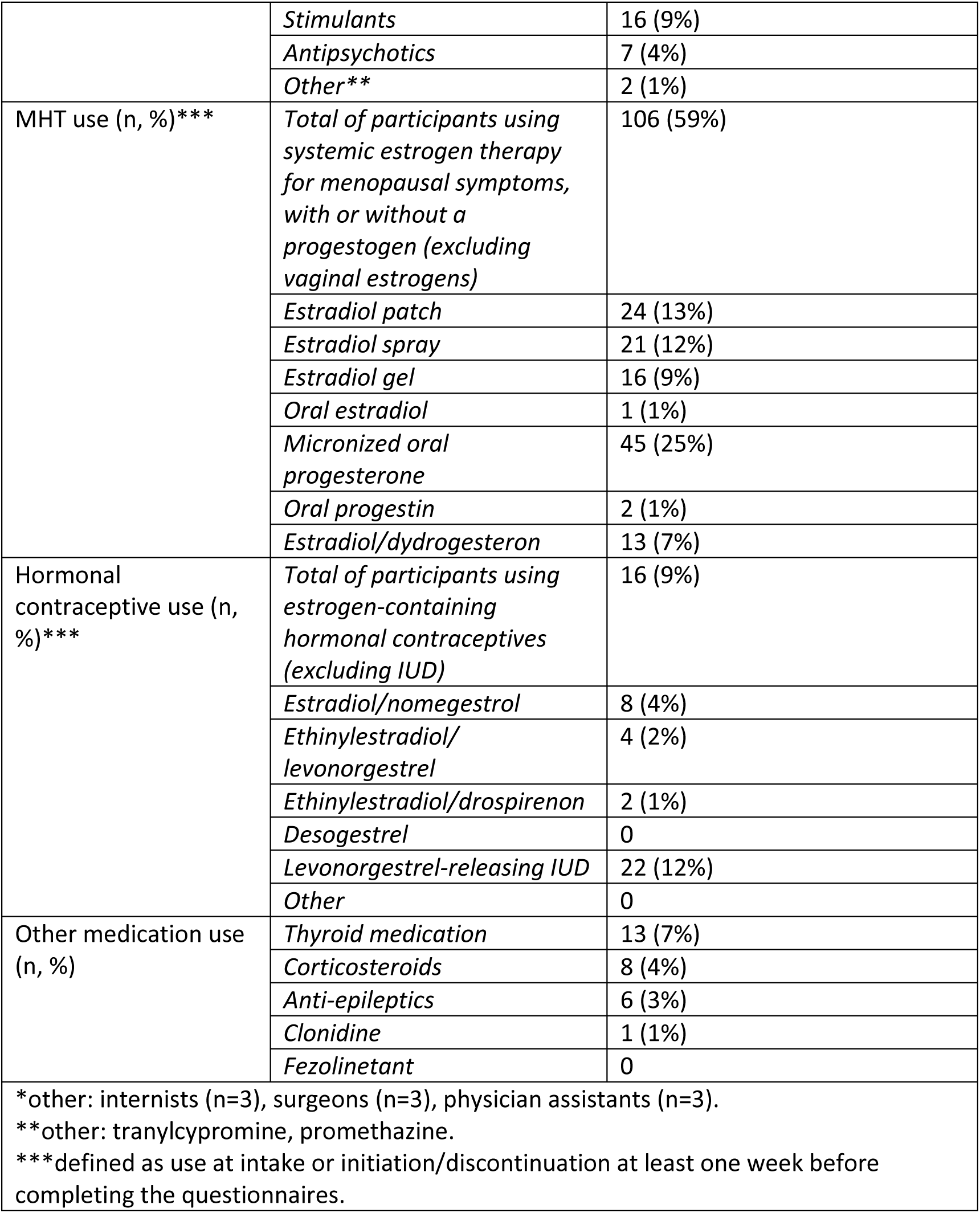
Demographic characteristics of the study participants.

| **Table 1.** Demographic characteristics of the study participants. |  |  |
| --- | --- | --- |
|  |  | **Total sample**<br>**N = 180** |
| **General information** |  |  |
| Age (median, IQR) |  | 51.0 (47.0–54.0) |
| Menopausal stage (n, %) | *Perimenopausal* | 110 (61%) |
|  | *Postmenopausal* | 70 (39%) |
| Referring physician (n, %) | *General practitioner* | 142 (79%) |
|  | *Gynecologist* | 24 (13%) |
|  | *Psychiatrist* | 4 (2%) |
|  | *Other\** | 9 (5%) |
| Type of menopause clinic at inclusion (n, %) | *Standard* | 157 (57%) |
|  | *Multidisciplinary* | 23 (13%) |
| Live births of pregnancy (n, %) | *0* | 58 (32%) |
|  | *1* | 34 (19%) |
|  | *2* | 60 (33%) |
|  | *3* | 22 (12%) |
|  | *≥4* | 6 (4%) |
| **Lifestyle factors** |  |  |
| Alcohol use (median, IQR) | *Consumptions per week* | 1.0 (0–4.0) |
|  | *Missing* | 3 (2%) |
| Drug use (n, %) | *No drug use* | 164 (91%) |
|  | *Drug use* | 7 (4%) |
|  | *Missing* | 9 (5%) |
| Smoking (n, %) | *No smoking* | 160 (89%) |
|  | *Smoking* | 14 (8%) |
|  | *Missing* | 6 (3%) |
| BMI (median, IQR) |  | 23.6 (21.5–27.4) |
|  | *Missing* | 15 (8%) |
| **Medication use** |  |  |
| Psychotropic medication use (n, %) | *Total of participants using at least one type of psychotropic medication* | 76 (42%) |
|  | *Antidepressants* | 49 (27%) |
|  | *Benzodiazepines* | 36 (20%) |
|  | *Stimulants* | 16 (9%) |
|  | *Antipsychotics* | 7 (4%) |
|  | *Other\*\** | 2 (1%) |
| MHT use (n, %)** | *Total of participants using systemic estrogen therapy for menopausal symptoms, with or without a progestogen (excluding vaginal estrogens)* | 106 (59%) |
|  | *Estradiol patch* | 24 (13%) |
|  | *Estradiol spray* | 21 (12%) |
|  | *Estradiol gel* | 16 (9%) |
|  | *Oral estradiol* | 1 (1%) |
|  | *Micronized oral progesterone* | 45 (25%) |
|  | *Oral progestin* | 2 (1%) |
|  | *Estradiol/dydrogesteron* | 13 (7%) |
| Hormonal contraceptive use (n, %)** | *Total of participants using estrogen-containing hormonal contraceptives (excluding IUD)* | 16 (9%) |
|  | *Estradiol/nomegestrol* | 8 (4%) |
|  | *Ethinylestradiol/levonorgestrel* | 4 (2%) |
|  | *Ethinylestradiol/drospirenon* | 2 (1%) |
|  | *Desogestrel* | 0 |
|  | *Levonorgestrel-releasing IUD* | 22 (12%) |
|  | *Other* | 0 |
| Other medication use (n, %) | *Thyroid medication* | 13 (7%) |
|  | *Corticosteroids* | 8 (4%) |
|  | *Anti-epileptics* | 6 (3%) |
|  | *Clonidine* | 1 (1%) |
|  | *Fezolinetant* | 0 |
| *other: internists (n=3), surgeons (n=3), physician assistants (n=3).<br>**other: tranlycypromine, promethazine.<br>***defined as use at intake or initiation/discontinuation at least one week before completing the questionnaires. |  |  |

### History of reproductive and non-reproductive depression

Premenstrual mood disorder was reported by 117 participants (65%), including 61 with moderate/severe PMS (52%) and 56 with PMDD (48%). Among parous participants, 72 reported perinatal depression (59%); 46 prenatal depression (64%), 61 postpartum depression (85%), and 35 both (49%). Non-reproductive depression was reported by 58 (32%) (Table 2).

**Table 2.** History of reproductive and non-reproductive depression.

| **Table 2.** History of reproductive and non-reproductive depression. |  |  |  |  |
| --- | --- | --- | --- | --- |
| **Premenstrual mood disorder (n, %)**<br>*PSST*<br>Sample: n = 180 | *No/mild PMS* | 63 (35%) |  |  |
|  | *Premenstrual mood disorder* | 117 (65%) | *Moderate/severe PMS* | 61 (52%) |
|  |  |  | *PMDD* | 56 (48%) |
| **Perinatal depression (n, %)**<br>*EPDS-Lifetime*<br>Sample: n = 122* | *No perinatal depression* | 50 (41%) |  |  |
|  | *Perinatal depression* | 72 (59%) | *Prenatal depression* | 46 (64%) |
|  |  |  | *Postpartum depression* | 61 (85%) |
| **Non-reproductive depression (n, %)**<br>*Clinical self-report*<br>Sample: n = 180 | *No depression* | 122 (68%) |  |  |
|  | *Non-reproductive depression* | 58 (32%) | *Confirmed diagnosis* | 48 (27%) |
|  |  |  | *Clinically suspected\*\** | 10 (6%) |
| *n = 58 (32%) had never had live births (parity = 0).<br>**Participants without a recorded diagnosis but with strong and consistent clinical indicators – such as long-term antidepressant use for mood symptoms, psychotherapy for mood symptoms, or other relevant treatments – were classified as having probable depression. All classifications were reviewed and confirmed by a psychiatrist to ensure clinical accuracy. |  |  |  |  |

### Associations between prior depression and current depressive symptom severity

Mean IDS-SR score was 28.5 (SD 12.5). Scores were higher in participants with premenstrual mood disorder (31.8, SD = 11.8), perinatal depression (30.7, SD = 11.5), and prior non-reproductive depression (31.4, SD = 12.7). Because these groups partly overlap, these values should be interpreted descriptively.

Table 3 and Figure 2 present the associations between prior depression and depressive symptom severity during the menopausal transition. In univariable models, all three exposures were significantly associated with higher IDS-SR scores (premenstrual mood disorder: B = 9.0, 95% CI 5.1–12.9, p < 0.001; perinatal depression: B = 7.8, 95% CI 3.4–12.1, p < 0.001; non-reproductive depression: B = 4.7, 95% CI 0.7–8.7, p = 0.021).

**Figure 2.**
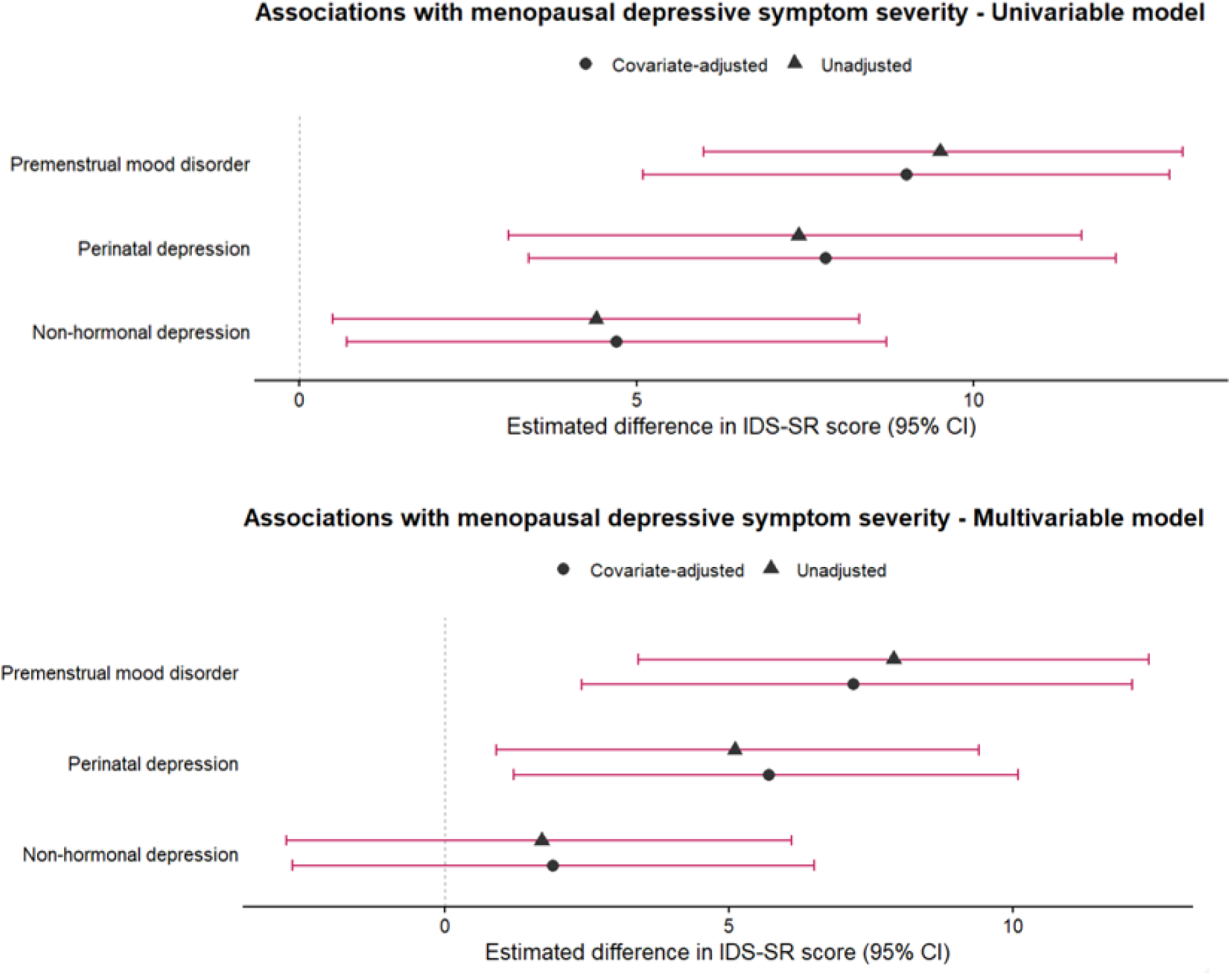
Estimated differences in IDS-SR scores during the menopausal transition associated with prior reproductive and non-reproductive depression.

**Table 3.** Associations between prior reproductive and non-reproductive depression, and severity of depressive symptoms (IDS-SR) during the menopausal transition.

|  |  |  |  |  |
| --- | --- | --- | --- | --- |
|  | B | 95% CI | P value | Model fit |
| Premenstrual mood disorder |  |  |  |  |
| N = 180 |  |  |  |  |
| Unadjusted | 9.5 | 6.0 to 13.1 | <0.001 | Log-likelihood: -696<br>R2: 0.13 |
| Covariate-adjusted* | 9.0 | 5.1 to 12.9 | <0.001 | Log-likelihood: -685<br>R2: 0.14 |
| Perinatal depression |  |  |  |  |
| N = 122 |  |  |  |  |
| Unadjusted | 7.4 | 3.1 to 11.6 | <0.001 | Log-likelihood: -471<br>R2: 0.09 |
| Covariate-adjusted* | 7.8 | 3.4 to 12.1 | <0.001 | Log-likelihood: -461<br>R2: 0.13 |
| Non-reproductive depression |  |  |  |  |
| N = 180 |  |  |  |  |
| Unadjusted | 4.4 | 0.5 to 8.3 | 0.027 | Log-likelihood: -706<br>R2: 0.03 |
| Covariate-adjusted* | 4.7 | 0.7 to 8.7 | 0.021 | Log-likelihood: -692<br>R2: 0.06 |
| Multivariable model |  |  |  |  |
| N = 122 |  |  |  |  |
| Premenstrual mood disorder | 7.9 | 3.4 to 12.4 | <0.001 | Log-likelihood: -465<br>R2: 0.18 |
| Perinatal depression | 5.1 | 0.9 to 9.4 | 0.019 |  |
| Non-reproductive depression | 1.7 | -2.8 to 6.1 | 0.46 |  |
| Multivariable covariate-adjusted* model |  |  |  |  |
| N = 120 |  |  |  |  |
| Premenstrual mood disorder | 7.2 | 2.4 to 12.1 | 0.0037 | Log-likelihood: -456<br>R2: 0.19 |
| Perinatal depression | 5.7 | 1.2 to 10.1 | 0.013 |  |
| Non-reproductive depression | 1.9 | -2.7 to 6.5 | 0.41 |  |
| *Adjusted for: age, MHT, oral contraceptives, smoking, alcohol. |  |  |  |  |

In the multivariable model including all three exposures (restricted to only parous participants, n=122), premenstrual mood disorder (B = 7.2, 95% CI 2.4–12.1, p = 0.0037) and perinatal depression (B = 5.7, 95% CI 1.2–10.1, p = 0.013) remained independently associated with higher IDS-SR scores, whereas prior non-reproductive depression was no longer significantly associated with IDS-SR scores (B = 1.9, 95% CI -2.7–6.5, p = 0.41). This model showed the best fit (log-likelihood = -456; R² = 0.19).

### Sensitivity analyses

Logistic regression models assessing probable current depression (IDS-SR ≥ 26) showed higher odds of depression among participants with premenstrual mood disorder (OR 4.0, 95% CI 2.0–8.1, p < 0.001) or perinatal depression (OR 3.6, 95% CI 1.6–7.9, p = 0.0015), but not in those with prior non-reproductive depression (OR 1.6, 95% CI 0.8–3.2, p = 0.17). Multivariable covariate-adjusted models showed similar findings (premenstrual mood disorder: OR 3.1, 95% CI 1.3–8.1, p = 0.014; perinatal depression: OR 2.7, 95% CI 1.2–6.3, p = 0.017; non-reproductive depression: OR 1.3, 95% CI 0.5–3.2, p = 0.55) (Supplementary table 1 and Supplementary figure 1).

In analyses restricted to perimenopausal participants, premenstrual mood disorder (B = 8.4, 95% CI 3.0–13.9, p = 0.0028) and perinatal depression (B = 6.6, 95% CI 1.5–11.7, p = 0.012) remained significantly associated with depressive symptom severity, whereas prior non-reproductive depression was not (B = 3.5, 95% CI -1.7–8.7, p = 0.19). Findings were similar in the multivariable model (premenstrual mood disorder: B = 7.1, 95% CI 1.1–13.1, p = 0.022; perinatal depression: B = 5.2, 95% CI 0.1–10.4, p = 0.048; non-reproductive depression: B = -0.5, 95% CI -6.1–5.0, p = 0.85) (Supplementary table 2).

When we used postpartum depression instead of perinatal depression as the exposure, results remained similar. Postpartum depression was significantly associated with depressive symptom severity (B = 6.7, 95% CI 2.4–11.0, p = 0.0026) and findings were consistent in the multivariable model (premenstrual mood disorder: B = 7.8, 95% CI 3.0–12.6, p = 0.0017; postpartum depression: B = 4.9, 95% CI 0.5–9.2, p = 0.028; non-reproductive depression: B = 1.9, 95% CI -2.7–6.5, p = 0.41) (Supplementary table 3).

## Discussion

This study aimed to examine whether prior reproductive and non-reproductive depression were differently associated with depressive symptom severity during the menopausal transition in a cohort of perimenopausal and postmenopausal individuals attending menopause outpatient clinics. Premenstrual mood disorder showed the strongest association with current depressive symptom severity, followed by perinatal depression and prior non-reproductive depression. After adjustment for overlap between exposures, premenstrual mood disorder and perinatal depression remained significantly associated with depressive symptom severity, whereas prior non-reproductive depression was no longer independently significantly associated. These findings suggest that prior reproductive depression may be more informative than a general history of depression for identifying vulnerability to depressive symptoms during the menopausal transition.

A possible explanation for our findings is that a subgroup of individuals shows heightened sensitivity to female reproductive hormone fluctuations across the life course, including during the premenstrual, perinatal, and perimenopausal phases. This concept has been described as hormonal mood sensitivity, a transdiagnostic vulnerability that may link PMDD, postpartum depression, and perimenopausal depression.^11–14^ Reproductive hormone changes may affect mood by modulating glutamatergic, GABAergic, dopaminergic, and serotonergic pathways.^9,10,14–16,45^ Progesterone-derived neuroactive metabolites, particularly allopregnanolone, may further influence mood through changes in GABA-A receptor sensitivity.^15^ Declining estrogen levels may also increase stress sensitivity by disrupting interactions between the hypothalamic-pituitary-gonadal and hypothalamic-pituitary-adrenal axes.^45,46^ These neurobiological mechanisms may be especially relevant during perimenopause, when hormonal fluctuations are most pronounced, and postpartum, when hormonal withdrawal may resemble perimenopause more closely than pregnancy. Two sensitivity analyses supported this interpretation. First, when we restricted the analysis to perimenopausal participants, premenstrual mood disorder and perinatal depression remained associated with depressive symptom severity, whereas prior non-reproductive depression did not. Likewise, when we replaced perinatal depression with postpartum depression, the results were similar.

Although neurobiological studies find biological mechanisms connecting reproductive hormone fluctuations and mood changes, hormonal mood sensitivity is unlikely to be purely biological. Psychosocial factors, like early childhood adversity and certain personality traits, may also shape how women respond to hormonal changes. Early childhood adversity has been linked to higher odds of PMDD, postpartum depression, and perimenopausal depressive symptoms.^47–49^ Neuroticism and self-criticism may further increase vulnerability, partly through irritability and poorer emotion regulation.^50–53^ This pattern suggests that hormonal changes may trigger depressive symptoms most strongly in persons with both biological sensitivity *and* psychosocial vulnerability.

Our findings have implications for both theory and clinical practice. They support a transdiagnostic framework of hormonal mood sensitivity, in which premenstrual, perinatal, and perimenopausal depression may reflect a shared vulnerability to reproductive hormone fluctuations rather than entirely distinct conditions. Clinically, this suggests that a detailed reproductive mood history may be more informative than a general history of depression when assessing vulnerability to depressive symptoms during the menopausal transition. This may improve identification of hormone-sensitive individuals and support more proactive monitoring during the menopausal transition and possibilities for preventative interventions. Emerging evidence suggests that MHT may benefit depressive symptoms in selected women^40–42^, underscoring the need for closer integration of gynecological and psychiatric care.

A key strength of this study is the detailed assessment of lifetime depression and reproductive history, which allowed us to compare prior reproductive and non-reproductive depression within the same cohort while accounting for their overlap. To our knowledge, this approach has not been taken in previous research. Sensitivity analyses supported the findings across alternative models, although subgroup results should be interpreted cautiously because of smaller sample sizes. The use of validated measures with good internal consistency further strengthens the robustness of the findings.

There are also several limitations to this study. First, we included women attending specialized menopause outpatient clinics, which may limit generalizability to community samples, although it increases relevance for specialized menopause care. Second, the cross-sectional design and retrospective assessment of prior reproductive and non-reproductive depression limit causal inference and introduce potential recall bias. Premenstrual mood disorders were assessed retrospectively using the PSST as a proxy for lifetime susceptibility. However, the PSST may overestimate PMDD and lacks specificity compared to prospective daily ratings.^54^ Similarly, the EPDS-Lifetime has been less extensively validated than the original EPDS. We also lacked information on prior hormonal contraceptive use and mood-related side effects. This may have introduced residual confounding, as hormonal contraceptive use has been associated with both lifetime depression risk and hormonally triggered mood disturbances.^43,55^ Additionally, we did not assess vasomotor symptoms and sleep disturbances despite their frequent co-occurrence and bidirectional associations with depressive symptoms.^1,56–59^ Finally, unavailable contextual factors such as socio-economic status, ethnicity, education, and major life events, may also have contributed to residual confounding.^5,60^

Our findings open several directions for future research. Prospective cohort studies on associations between prior reproductive and non-reproductive depression and depressive symptoms during the menopausal transition should be conducted in community-based populations. They should also assess experiences with hormonal contraceptives as a potential marker of hormonal mood sensitivity, as well as other vulnerability factors such as early life adversity and personality traits.

Future work should also test whether hormone-sensitive and non-hormone-sensitive subgroups differ in their clinical presentation, as reproductive depressions may involve more symptoms like mood lability, irritability, anxiety, and sensitivity to interpersonal stress than non-reproductive depression.^35,61–66^ Prospective studies across the reproductive lifespan should then examine whether these subgroups differ in symptom course and in their response to hormonal and non-hormonal treatments during the menopausal transition.

In conclusion, we found that prior reproductive depression, including premenstrual mood disorder and perinatal depression, but not prior non-reproductive depression, was associated with greater depressive symptom severity during the menopausal transition. This pattern supports the concept of a hormonally sensitive subgroup of individuals who are especially vulnerable to mood disturbances during female reproductive transitions. Recognizing hormonal mood sensitivity could improve early identification of individuals at risk and support more targeted and effective interventions for depression in midlife.

## Data Availability

Data in the current study are available upon reasonable request to the authors.

## CRediT statement

**MS:** Conceptualization, Methodology, Formal analysis, Investigation, Data Curation, Writing – Original Draft, Visualization. **MWLM:** Conceptualization, Methodology, Formal analysis, Data Visualization, Writing – Review & Editing. **DKED:** Conceptualization, Resources, Writing – Review & Editing. **YR:** Conceptualization, Resources, Writing – Review & Editing. **BFPB:** Conceptualization, Methodology, Writing – Review & Editing, Supervision, Funding Acquisition.

All authors agree to be accountable for all aspects of the work.

## Acknowledgements

We thank all participants of the MOPP study and the healthcare professionals involved in recruitment for their crucial contributions to this research.

