## Supplementary materials for "Prior reproductive and non-reproductive depression, and depressive symptoms in menopausal transition"

**Supplementary table 1.** Odds ratios for depression during the menopausal transition, defined as IDS-SR ≥ 26 (n=108, 60%), according to prior reproductive and non-reproductive depression.

| | OR | 95% CI | P value | Model fit |
|---|---|---|---|---|
| **Premenstrual mood disorder**<br>N = 180 | | | | |
| Unadjusted | 4.1 | 2.2 to 8.0 | <0.001 | Log-like: -111 |
| Covariate-adjusted* | 4.0 | 2.0 to 8.1 | <0.001 | Log-like: -110 |
| **Perinatal depression**<br>N = 122 | | | | |
| Unadjusted | 3.4 | 1.6 to 7.4 | 0.0015 | Log-like: -78 |
| Covariate-adjusted* | 3.6 | 1.6 to 7.9 | 0.0015 | Log-like: -75 |
| **Non-reproductive depression**<br>N = 180 | | | | |
| Unadjusted | 1.6 | 0.8 to 3.1 | 0.17 | Log-like: -120 |
| Covariate-adjusted* | 1.6 | 0.8 to 3.2 | 0.17 | Log-like: -116 |
| **Multivariable model**<br>N = 122 | | | | |
| Premenstrual mood disorder | 3.4 | 1.5 to 8.2 | 0.0049 | Log-like: -74 |
| Perinatal depression | 2.6 | 1.2 to 5.9 | 0.019 | |
| Non-reproductive depression | 1.3 | 0.5 to 3.1 | 0.56 | |
| **Multivariable covariate-adjusted* model**<br>N = 120 | | | | |
| Premenstrual mood disorder | 3.1 | 1.3 to 8.1 | 0.014 | Log-like: -72 |
| Perinatal depression | 2.7 | 1.2 to 6.3 | 0.017 | |
| Non-reproductive depression | 1.3 | 0.5 to 3.2 | 0.55 | |

*Adjusted for: age, MHT, oral contraceptives, smoking, alcohol.

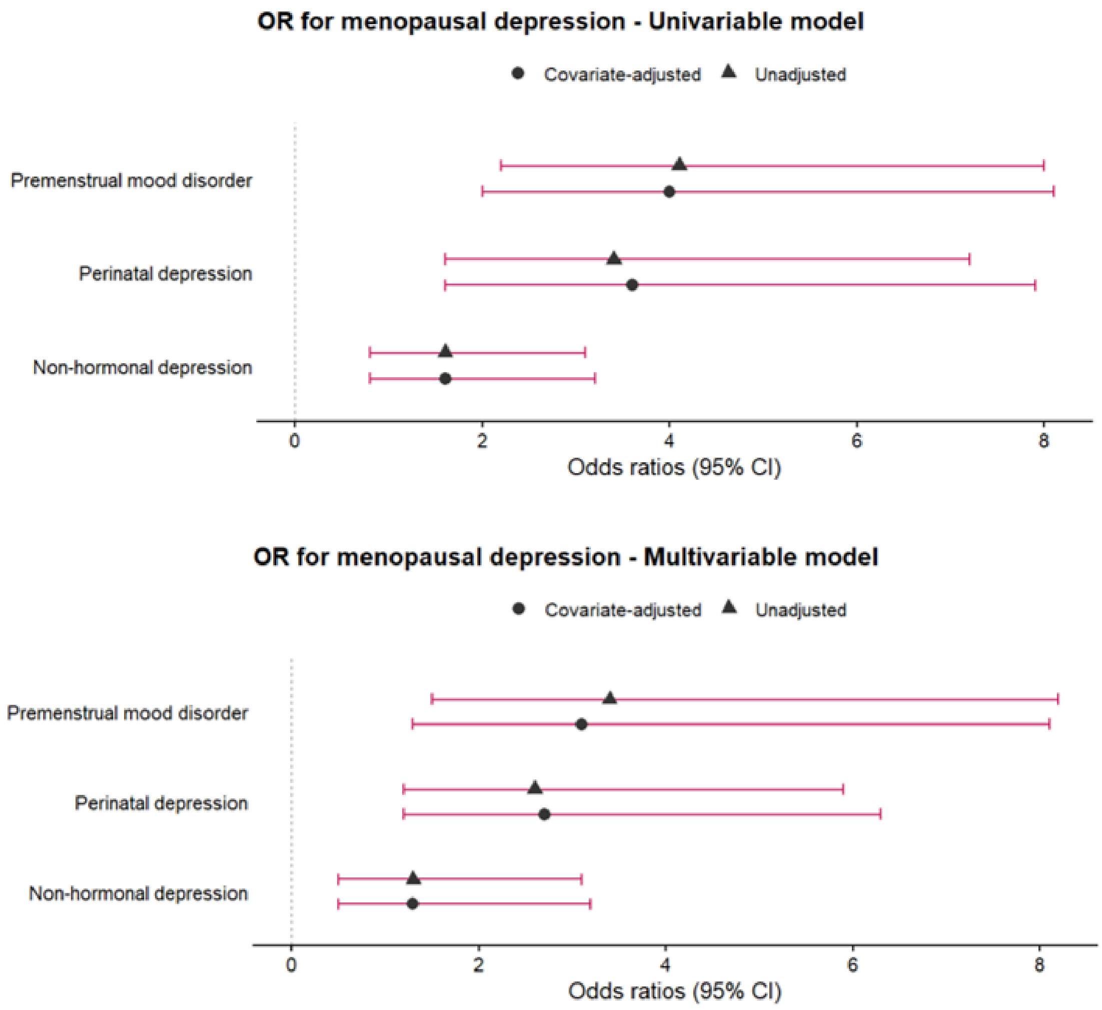


**Supplementary figure 1**. OR for depression during the menopausal transition.

**Supplementary table 2.** Associations between prior reproductive and non-reproductive depression, and severity of depressive symptoms (IDS-SR) during the menopausal transition among perimenopausal participants.

| | B | 95% CI | P value | Model fit |
|---|---|---|---|---|
| **Premenstrual mood disorder**<br>N = 110 | | | | |
| Unadjusted | 8.6 | 3.4 to 13.7 | 0.0013 | Log-like: -427<br>R2: 0.09 |
| Covariate-adjusted* | 8.4 | 3.0 to 13.9 | 0.0028 | Log-like: -420<br>R2: 0.10 |
| **Perinatal depression**<br>N = 79 | | | | |
| Unadjusted | 6.3 | 1.4 to 11.3 | 0.013 | Log-like: -301<br>R2: 0.08 |
| Covariate-adjusted* | 6.6 | 1.5 to 11.7 | 0.012 | Log-like: -295<br>R2: 0.13 |
| **Non-reproductive depression**<br>N = 110 | | | | |
| Unadjusted | 2.9 | -2.0 to 7.9 | 0.24 | Log-like: -432<br>R2: 0.01 |
| Covariate-adjusted* | 3.5 | -1.7 to 8.7 | 0.19 | Log-like: -423<br>R2: 0.03 |
| **Multivariable model**<br>N = 79 | | | | |
| Premenstrual mood disorder | 7.0 | 1.3 to 12.6 | 0.016 | Log-like: -297<br>R2: 0.15 |
| Perinatal depression | 5.1 | 0.0 to 10.1 | 0.049 | |
| Non-reproductive depression | -0.5 | -5.9 to 4.9 | 0.86 | |
| **Multivariable covariate-adjusted* model**<br>N = 79 | | | | |
| Premenstrual mood disorder | 7.1 | 1.1 to 13.1 | 0.022 | Log-like: -292<br>R2: 0.20 |
| Perinatal depression | 5.2 | 0.1 to 10.4 | 0.048 | |
| Non-reproductive depression | -0.5 | -6.1 to 5.0 | 0.85 | |

*Adjusted for: age, MHT, oral contraceptives, smoking, alcohol.

**Supplementary table 3.** Associations between prior reproductive and non-reproductive depression, and severity of depressive symptoms (IDS-SR) during the menopausal transition, using postpartum depression instead of peripartum depression as exposure.

| | B | 95% CI | P value | Model fit |
|---|---|---|---|---|
| **Postpartum depression**<br>N = 122 | | | | |
| Unadjusted | 6.4 | 2.2 to 10.6 | 0.0030 | Log-like: -472<br>R2: 0.07 |
| Covariate-adjusted* | 6.7 | 2.4 to 11.0 | 0.0026 | Log-like: -463<br>R2: 0.10 |
| **Multivariable model**<br>N = 122 | | | | |
| Premenstrual mood disorder | 8.3 | 3.9 to 12.8 | <0.001 | Log-like: -466<br>R2: 0.17 |
| Postpartum depression | 4.6 | 0.4 to 8.7 | 0.032 | |
| Non-reproductive depression | 1.7 | -2.7 to 6.2 | 0.44 | |
| **Multivariable covariate-adjusted* model**<br>N = 120 | | | | |
| Premenstrual mood disorder | 7.8 | 3.0 to 12.6 | 0.0017 | Log-like: -457<br>R2: 0.18 |
| Postpartum depression | 4.9 | 0.5 to 9.2 | 0.028 | |
| Non-reproductive depression | 1.9 | -2.7 to 6.5 | 0.41 | |

*Adjusted for: age, MHT, oral contraceptives, smoking, alcohol.
